# Knowledge, attitude and practices on intermittent preventive treatment in pregnant women with malaria: a mixed method facility-based study in Western Kenya

**DOI:** 10.1101/2023.10.20.23296986

**Authors:** Joseph Mukala, Dominic Mogere, Peter Kirira, Bernard N. Kanoi, Violet Akinyi, Francis Kobia, Harrison Waweru, Jesse Gitaka

## Abstract

Intermittent preventive treatment remains a core strategy for malaria prevention in pregnancy. Sulfadoxine-pyrimethamine is recommended for all pregnant women in malaria prone zones. It is scheduled monthly at each antenatal care visit up to 36 weeks. Here, we sought to assess the knowledge, attitude and practices on intermittent preventive treatment in pregnant women with malaria in Webuye hospital. Prior to the enrollment, ethical approval and permissions were sought from relevant institutions, as well as consents obtained from 140 participants aged between 18-49 years with gestation about 16 weeks. Malaria test was conducted via either microscopy or rapid test and participants were split into malaria positive and negative cohorts. Closed and open-ended questionnaire were administered to the participants and two focus group discussions were organized to collect their views. The results were expressed in percentage and Chi-square of association at a p-value equal or less than 0.05 (95%). Qualitative data were analyzed by the means of MAXQDA software. Our analysis revealed that there was a significant difference between the proportion of negative and positive groups among mothers’ knowledgeable on the side effects (p-value = 0.001), different doses (p-value = 0.012). Those who were informed about intermittent preventive treatment before administration (p-value = 0.003). The proportion of mothers knowledgeable about side effects and different doses were higher among the malaria positive group as compare to the negative with 52.9% versus 25.7% and 20.0% versus 5.7% respectively. Moreover, 76.3% of respondents reported that intermittent preventive treatment prevents malaria, 30.9% had the opinion that it causes abortion. Expectant women who were aware of the benefits of this strategy had this to say; “*This medicine helps to reduce the effects of malaria and prevents mother to contract malaria”.* However, those unaware had this to say; *“I have never been told about something like that but it prevents diseases”. Those who* knew about the schedule and side effects said *“It is given three doses during each antenatal visit”.* Therefore, good knowledge, attitude and practices of intermittent preventive treatment are key for control and prevention of malaria in endemic prone areas.

## Introduction

Globally, malaria infection is a serious communicable infectious disease that threatens the life of the half of world population with approximately 515 million people in Latin America, Asia and Sub-Saharan Africa region with one to three million deaths each year [1]. Recently, malaria has affected 228 million people worldwide, with approximately 213 million in the Sub-Saharan Africa representing 93% of the total population. The most vulnerable persons being children and expectant women with consequences ranging from deadly complications such as anemia, abortion, intrauterine fetal retardation, small gestation for age, prematurity and low birth weight. Indeed, intermittent preventive treatment is a strongly recommended molecule for all pregnant women in moderate to high malaria transmission zones except HIV patients, and Kenya adopted three doses which are efficient and efficacious to protect pregnancy against malaria [2, 3].

The utilization and uptake of highly cost-effective interventions of malaria were found associated with poor maternal knowledge, complicated guidelines and policies issues baring healthcare workers to deliver efficient routine antenatal care. However, different barriers to access these interventions are poorly investigated in the current context. Therefore, innovation in terms of malaria identification, eradication and diagnostic strategies are more needed [4]. Despite the recommendation for pregnant women living in endemic malaria zones to be receiving sulfadoxine-pyrimethamine, Kenya presented sub-optimal uptake of IPT-SP estimated at 13% of those who received three or more doses, while 28% received 1 or more doses. However, Bungoma County recorded 45% of pregnant women who received more than three doses [5].

Malaria strategy plan 2019-2023 had set its goal at 100% coverage of all people at risk dwelling in malaria prone zones through access to effective malaria preventive interventions. In Kenya, the increase of utilization was fixed at least at 80 percent by 2023[6]. WHO established a malaria free world vision stating that countries should take opportunities to innovate local and adapted activities lying with global recommendations such as equity in access to health services especially for the most vulnerable and hard-to-reach populations. Therefore, this goal translates into actions which can reduce malaria incidence and mortality gradually from 90% to 40% between 2020 up to 2030 [7]. A multiple indicator cluster survey carried out among pregnant women in Bungoma County, which is one of holo-endemic malaria areas in Western Kenya region, highlighted that out of 57% of population who slept under the mosquito nets, 70% of them were pregnant, 22% only had received 2 doses and plus of intermittent preventive treatment. In addition, The survey stated that there were 31 deaths among children of less than five years over 1000 live births against a recorded national average of 22 deaths over 1000 live births which should be addressed to fill the gap [8],[9],[10]. The County Government of Bungoma listed malaria and anemia as the most frequent disease burden in its integrated development plan 2018-22 and targeted to bridge existing gap by improving the low rate of maternal and infant indicators through the offer of quality antenatal care service to the vulnerable population. It was established that the half of pregnant women in the county had not achieved ANC four visits as recommended by the WHO guideline [11]. Roll Back Malaria partnership emphasizes that the antenatal care should be an entry point where every pregnant woman will be served with at least one dose of Fansidar and at least 80% will receive insecticide treated nets with ambition to reach 100% by 2025 [12].

Previous studies have showed benefits associated with the use of intermittent preventive treatment with regard to knowledge of expectant women, but only few have put accent on what pregnant women knew and translated into actions “practices”. The explicit use of qualitative opinions and views of main beneficiaries on IPT/SP benefits, side effects, schedule, doses, safety, prior information, attitude of healthcare providers, trust and sources of information can improve the uptake and the utilization of intermittent preventive treatment.

## Materials and methods

### Study design

A facility mixed methods study design comprising qualitative-quantitative parameters drawn from a prospective cohort research study conducted at Webuye hospital from (March 2022) to (December 2022). Participants were enrolled from 16 weeks of gestation and followed-up to the delivery. The study site has been extensively discussed elsewhere [13],[14].

### Sampling frame and inclusion criteria

In this study, a total of 140 pregnant women aged between 18-49 years with gestation at about 16 weeks were selected from the ANC clinic. To be eligible for the study, participants needed to be mentally stable and residents in the area for almost six months. Malaria testing was conducted using either microscopy or rapid diagnostic tests. Out of the 140 participants, 70 (50%) tested positive for malaria while the other tested negative.

### Collection of data

Data were collected using the following steps: The questionnaire was pretested and administered to participants in English, with Kiswahili translations provided for those with language barriers. Questions were appropriately formulated, numbered, and provided with options for both close-ended and open-ended answers. They were also scored, coded, and validated. Before enrollment, participants were informed about the consent process, including subject respects, the rights to participate or to withdraw at any time, benefits, confidentiality, compensation, and the voluntary nature of participation. Participants were also informed about the privacy of video and image recording during the focus group discussions. Questions related to the benefits, side effects, doses, schedule, safety, healthcare worker attitudes during administration of IPT-SP, prior information on the medicine, and other sources of information were included in the questionnaire. Two focus group discussions were organized, lasting for 60 minutes each. The research team performed video and image recording, took notes, and kept track of timing.

### Ethical considerations

Ethical approval was sought from the Ethics Review Committee of Mount Kenya University, and a research permit obtained from NACOSTI (MKU/ERC/2100, license No. NACOSTI/P/22/16233), as well as the local authorizations from County and Webuye hospital authorities. Participants were explained on the confidentiality, voluntarily participation, respect, withdrawal, compensation and agreement were obtained before video tape recording and images which were to be discarded immediately after the study.

### Data analysis

The sample size calculation formula for cohort was used on the basis of malaria prevalence in the non-exposed group estimated at 28% according to the study of Nyamu [15]. The prevalence of malaria in the exposed group was estimated at 6.1% according to the DHIS2 [16]. Beta (10%), Alpha (5%), Confidence level of 95%, Z alpha (1.96), Z beta value (1.28), Sample size for group-1(n1=60), Sample size for each group-2 (n2=60), Sample size for both group (n1+n2=120), Attrition (=20%), Total sample size with attrition=144. Answers were fed into SPSS 27, eight questions were administered to the participants split into positive and negative malaria cohorts, and answers were noted. The results were presented in percentage under the tables and figures. Themes and sub-themes were developed during two focus group discussions and captured under the form of in-depth views and opinions among positive and negative malaria cohorts, as well as video recording, transcribing, coding and analyzing using MAXQDA software. Chi-square test of association was computed to test statistical significance at p-value equal or less than 0.05 (95%).

## Results

There was a significant difference in proportion between negative and positive malaria groups among mothers’ knowledgeable on the side effects (p-value = 0.001), different doses (p-value = 0.012), and those who were informed about intermittent preventive treatment before administration (p-value = 0.003), The proportion of mothers knowledgeable about side effects and different doses were higher among the malaria positive group as compare to the negative group (52.9% versus 25.7% and 20.0% versus 5.7%) respectively. However, only half of the malaria positive group were informed about intermittent preventive treatment sulfadoxine-pyrimethamine before it was administered as compare to 74.3% in the negative group. Majority of pregnant women 70 (49.9%) reported that healthcare workers (HCW) had good attitude towards provision of the service. 124 (88.6%) respondents trusted the information given by healthcare workers. [Table 1].

**Table 1:** Intermittent Preventive Treatment.

| Variables | Malaria test |  |  | p-value |
| --- | --- | --- | --- | --- |
|  | Overall,<br>N = 140 | Negative,<br>n = 70 | Positive,<br>n = 70 |  |
| Benefits of IPT, n (%) |  |  |  | 0.610 |
| No | 63 (45.0) | 33 (47.1) | 30 (42.9) |  |
| Yes | 77 (55.0) | 37 (52.9) | 40 (57.1) |  |
| Knowledge of side effects of IPT, n (%) | 55 (39.3) | 18 (25.7) | 37 (52.9) | 0.001 |
| Different doses of IPT, n (%) | 18 (12.9) | 4 (5.7) | 14 (20.0) | 0.012 |
| Safe drug during first trimester, n (%) | 83 (59.3) | 47 (67.1) | 36 (51.4) | 0.058 |
| Ever refused to take IPT, n (%) | 20 (14.3) | 13 (18.6) | 7 (10.0) | 0.150 |
| Have you been informed about IPT, n (%) | 87 (62.1) | 52 (74.3) | 35 (50.0) | 0.003 |
| Trust information given by HCW, n (%) | 124 (88.6) | 65 (92.9) | 59 (84.3) | 0.110 |
| Attitude of HCW towards provision, n (%) |  |  |  | 0.300 |
| Bad | 24 (17.1) | 9 (12.9) | 15 (21.4) |  |
| Good | 57 (40.7) | 32 (45.7) | 25 (35.7) |  |
| Very good | 70 (49.9) | 34 (48.5) | 36 (51.4) |  |
| IPT Doses given, n (%) |  |  |  | <0.001 |
| None | 29 (20.7) | 24 (34.3) | 5 (7.1) |  |
| Once | 34 (24.3) | 17 (24.3) | 17 (24.3) |  |
| Twice | 46 (32.9) | 11 (15.7) | 35 (50.0) |  |
| Thrice | 31 (22.1) | 18 (25.7) | 13 (18.6) |  |

more than half of respondents either 7(58%) were aware of the benefit of sulfadoxine-pyrimethamine against 5 (42%). Pregnant women expressed that Sulfadoxine-pyrimethamine prevents malaria and those aware were 7 (58%) [Table 2].

**Table 2:** Benefits of Sulfadoxine-pyrimethamine.

| Knowledge of the benefit of IPT-SP | Frequency | Percentage | Cumulative |
| --- | --- | --- | --- |
| Aware | 7 | 58% | 58% |
| Unaware | 5 | 42% | 100% |
| Total | 12 | 100 % | 100% |

Percentage was used to calculate knowledge awareness among participants.

There was statistical association between the number of IPT-SP doses and malaria test (p-value <0.001). Overall, most women received two doses of IPT (n = 46; 32.9%). Among the malaria negative cohort, 24 (34.3%) did not receive any dose while 11 (15.7%) received two doses. This was different with malaria positive cohort 35 (50.0%) who received two doses of IPT-SP and only 5 (7.1%) had received any dose. The total coverage of 2 doses and + was higher among the positive pregnant women 48 (68.6%) against negative pregnant women 29 (41.4%) [Table 3].

**Table 3:** Different Doses of IPT Received by Pregnant Women.

| Variables | Malaria test |  |  | P-value |
| --- | --- | --- | --- | --- |
|  | Overall,<br>N = 140 | Negative,<br>N = 70 | Positive,<br>N = 70 |  |
| **IPT Doses given, n (%)** |  |  |  | <.0001 |
| None | 29 (20.7) | 24 (34.3) | 5 (7.1) |  |
| Once | 34 (24.3) | 17 (24.3) | 17 (24.3) |  |
| Twice | 46 (32.9) | 11 (15.7) | 35 (50.0) |  |
| Thrice | 31 (22.1) | 18 (25.7) | 13 (18.6) |  |

Focus group discussions were organized to collect pregnant women opinions as well as their views, which were captured, recorded, transcribed, then coded and analyzed by the mean of MAXQDA Software analysis to identify practices with regard to the completeness of the sulfadoxine-pyrimethamine during pregnancy. The results showed that 10 (83%) pregnant women had completed IPT/SP doses which is considered as a key element toward malaria prevention during pregnancy [Table 4].

**Table 4:** Practices of intermittent preventive treatment.

| Sulfadoxine-pyrimethamine | Frequency | Percentage |
| --- | --- | --- |
| Practicing/completing | 10 | 83% |
| No practicing/refusing | 2 | 17% |
| **Total** | **12** | **100%** |

Views of respondents expressed in percentage with regard to the completeness of IPT-SP.

The benefits of using IPT-SP during pregnancy among the 140 women interviewed, 75 (76.3%) reported that IPT-SP prevents malaria, followed by 24 (13.2%) who reported that it prevents malaria and protects baby and mother [Figure 1].

**Figure 1:**
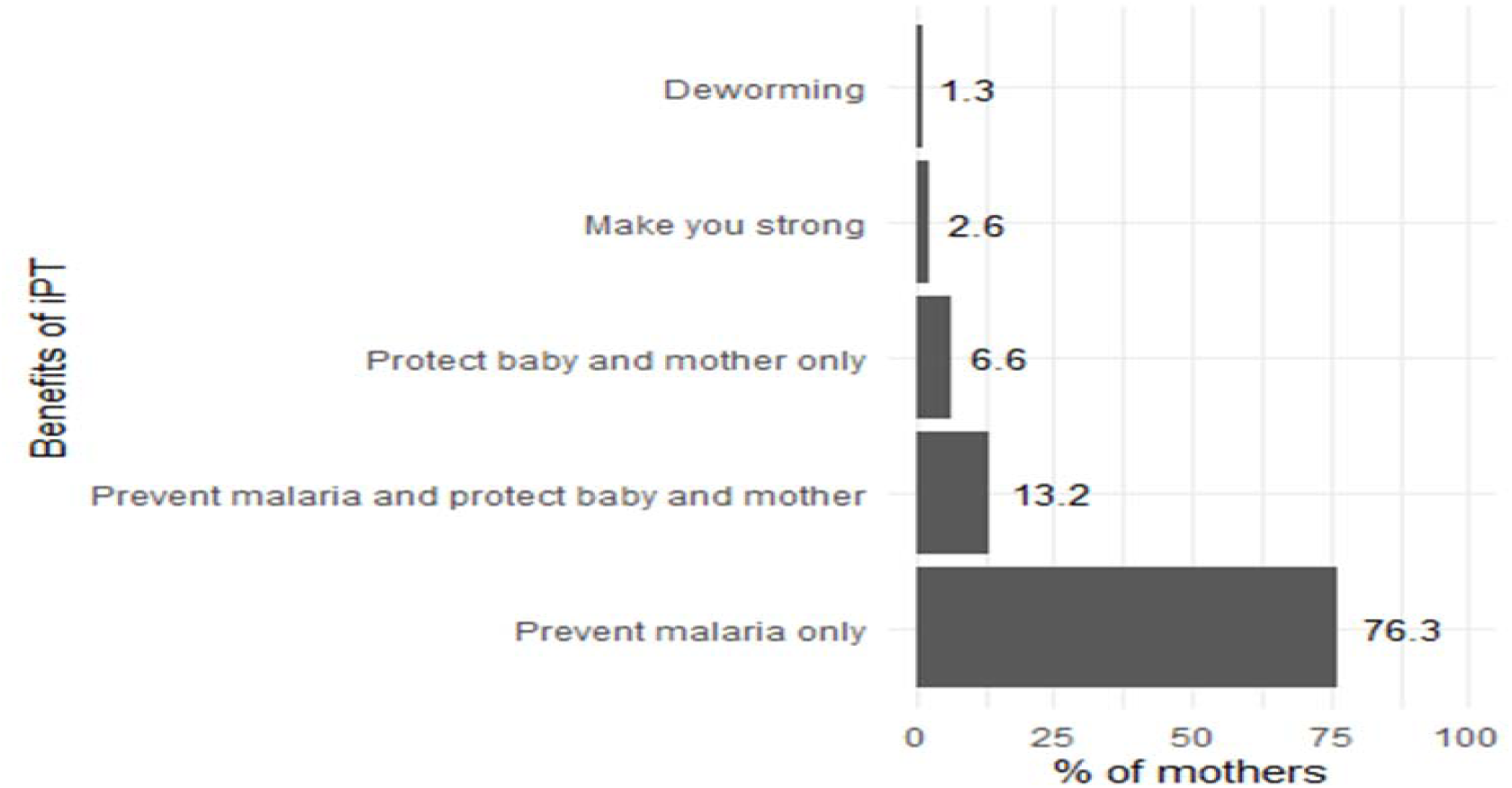
Benefits of intermittent preventive treatment. Percentage illustration on the benefits of IPT-SP.

The study also assessed the reason why it is unsafe to use IPT-SP during the first trimester. Among the 140 participants interviewed, 30.9% had the opinion that it causes abortion, 10.9% said it causes vomiting, 9.1% premature labor and 7.3% said it causes fatigue as well as other reasons [Figure 2].

**Figure 2:**
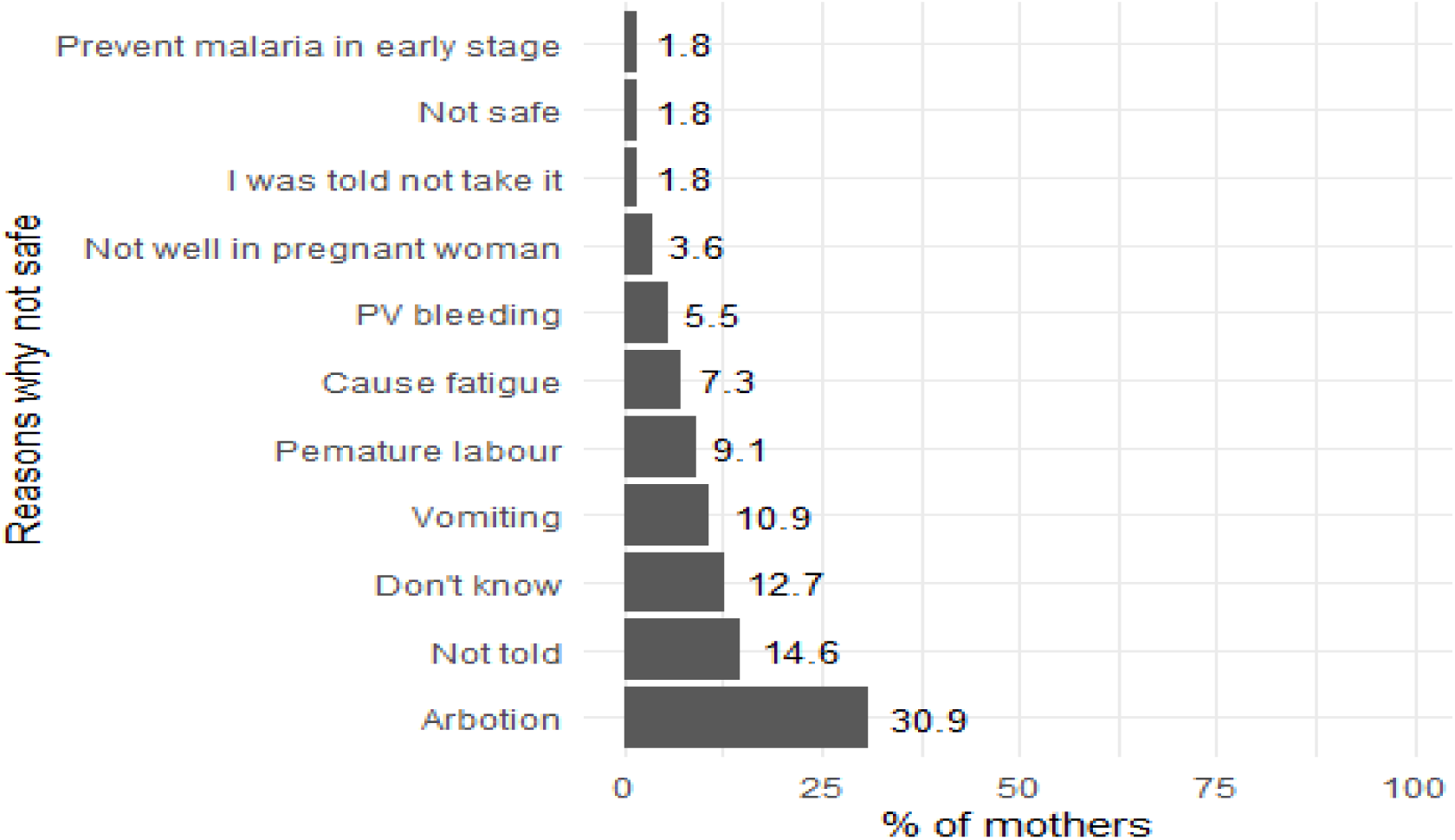
Reasons of Not Using IPT-SP during the First Trimester. Percentage illustration of reasons evoked for not using IPT-SP

There was 65 (92%) coverage of IPT/SP 2+ doses among positive malaria women against 46 (55.7%) among negative malaria [Supplementary Figure 3]

The cohort of malaria positive respondents 35 (50%) received 2 doses of IPT-SP against 11 (15.7%) in the cohort of malaria negative. Those who received 3 doses were 18 (25.7%) in negative cohort versus 13 (18.6%) in positive cohort [Supplementary Figure 4].

Practices of intermittent preventive treatment among pregnant women with malaria, Sub-theme: Acceptance and completeness of sulfadoxine-pyrimethamine: “*I swallowed this medicine twice or thrice and did not refuse it*” [Supplementary Figure 5].

## Discussion

In a research survey done in Sabatia Kenya under a cross-sectional design, it was found that a good number of pregnant women had good knowledge of intermittent preventive treatment sulfadoxine-pyrimethamine benefits, but did not know the exact time for the beginning of intermittent preventive treatment/sulfadoxine-pyrimethamine and never experienced sulfadoxine-pyrimethamine side effects [17]. However, this study found that marital status, knowledge of benefits of sulfadoxine-pyrimethamine and gestation age were significantly associated with uptake of sulfadoxine-pyrimethamine, with women who had good knowledge of benefits having higher likelihood of receiving the third dose than those with poor knowledge. In our study, we found that the two key variables which determined completeness of IPT/SP were practices and malaria test. The first was found to have been associated with completeness through qualitative opinion in that pregnant women with good practices had this to say “I had finished 2 or 3 doses, whereas the second factor showed that pregnant women with positive malaria test had higher likelihood to complete the second dose as compare to those with negative malaria test.

A research survey under a qualitative research design conducted in two countries; Mali and Kenya using focus group discussion, found that having correct knowledge of doses and IPT/SP intake intervals, expectant women felt that the method was still very powerful to be avoided during pregnancy due to possible induction of miscarriage [19]. The result of our study found that 30% of participants had the same opinion that IPT/SP causes abortion. Strikingly, the safety variable as an important parameter on the side of pregnant women and their inborn infants was raised as a reason why pregnant women do not accept IPT/SP during the first trimester but also for the subsequent doses. Similarly the same concern was raised in the research study conducted in Somalia [23].

We found that the result of our study was in agreement with a qualitative study carried out in Mozambique citing the word of a pregnant women going to antenatal clinic who recognized sulfadoxine-pyrimethamine tablet as the white tablets given to pregnant women, and took in the presence of healthcare worker. Saying that *“I was not told more but I was given three tablets which I took”. Strikingly, she argued that “Some pregnant women even when advised and counselled do not complete the doses of sulfadoxine-pyrimethamine” (maternal and child nurse)* [20]. A descriptive research design found that approximately all respondents had heard about IPT/SP and 57% who had stated that this medicine was convenient with malaria prevention in both mothers and unborn children, and 15.4% felt that it was used to treat malaria. However, nausea, vomiting, body weakness, headache, dizziness, abdominal pain and diarrhea were reported as unwanted effects of the molecule used to prevent malaria in expectant women. This previous finding corroborates with the finding of our study which showed that these unwanted side effects did not prevent pregnant women to receive the subsequent doses of IPT/SP. Therefore, we found that knowledge of pregnant women vis-a-vis of IPT/SP benefit, schedule, doses and side effects were acceptable and can be translated into good practices as demonstrated in the qualitative insight. We are in support of the following studies which were carried out in Uganda and Nigeria. According to their findings, the Ugandan study laid down the credence that the attendance to antenatal clinic increased the accessibility to the IPT/SP, while the Nigerian study argued that all pregnant women do not attend the clinic session during pregnancy. Therefore, giving room to support the work of healthcare providers prior information given to pregnant women to empower them on IPT/SP, which can translate later into acceptance and completeness of this medicine used for malaria prevention, beyond that laying a good ground to complement community strategies to reach every pregnant woman [18], [21], [22], [25].

## Conclusion

Good knowledge and prior information by the patients on the side effects and dosage of intermittent preventive treatment, as well as patients’ attitudes and practices were key for efficient prevention of malaria in both cohorts. They should therefore be further leveraged to empower pregnant women in malaria endemic areas as an effective preventive strategy towards universal IPT-SP coverage for improved maternal and child outcomes.

## Data Availability

All data produced in the present study are available upon reasonable request to the author

---

**Supplementary figure 3:**
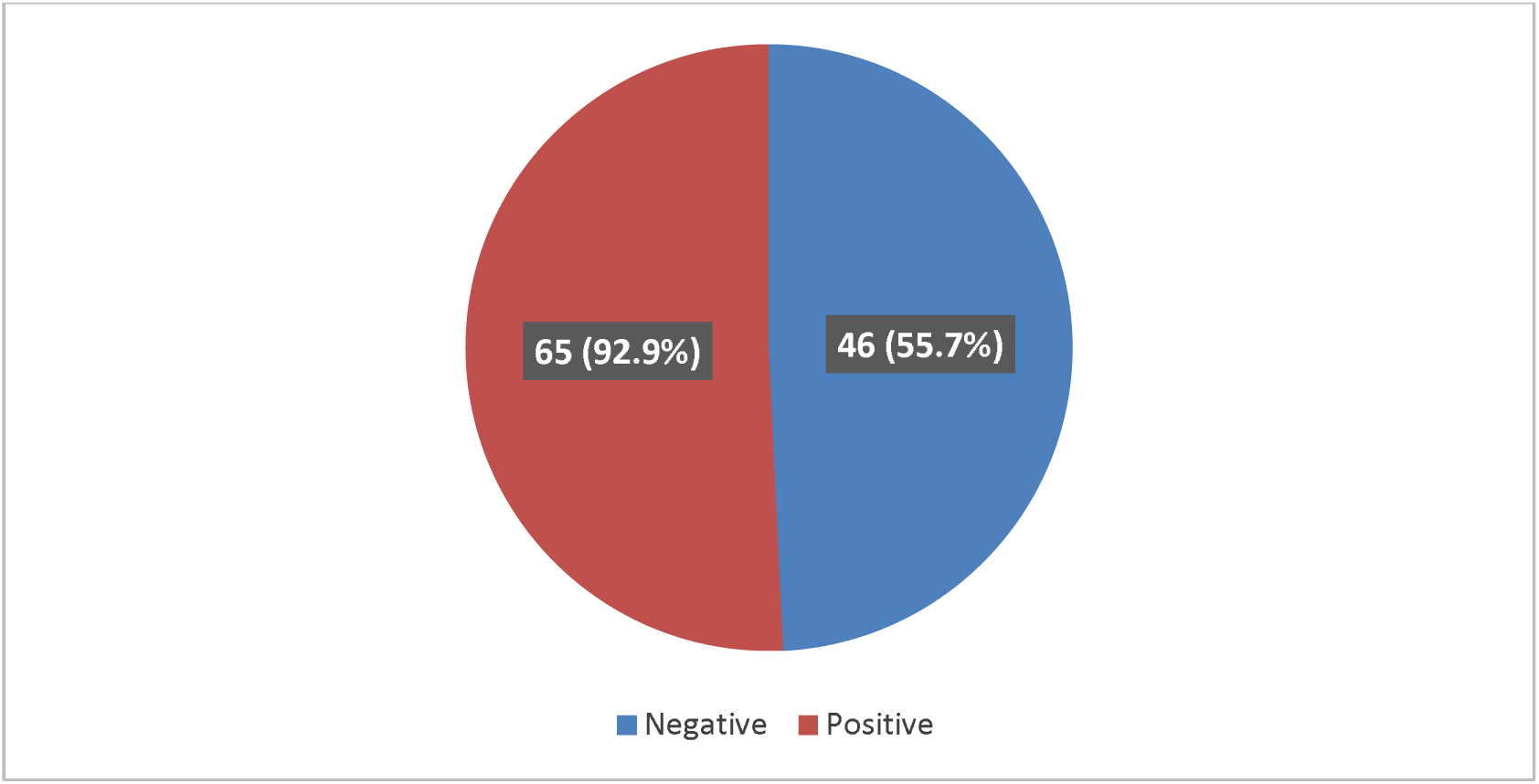
IPT-SP coverage and malaria test among pregnant women. Percentage illustration of IPT-SP coverage versus malaria test for above and less than two doses.

**Supplementary figure 4:**
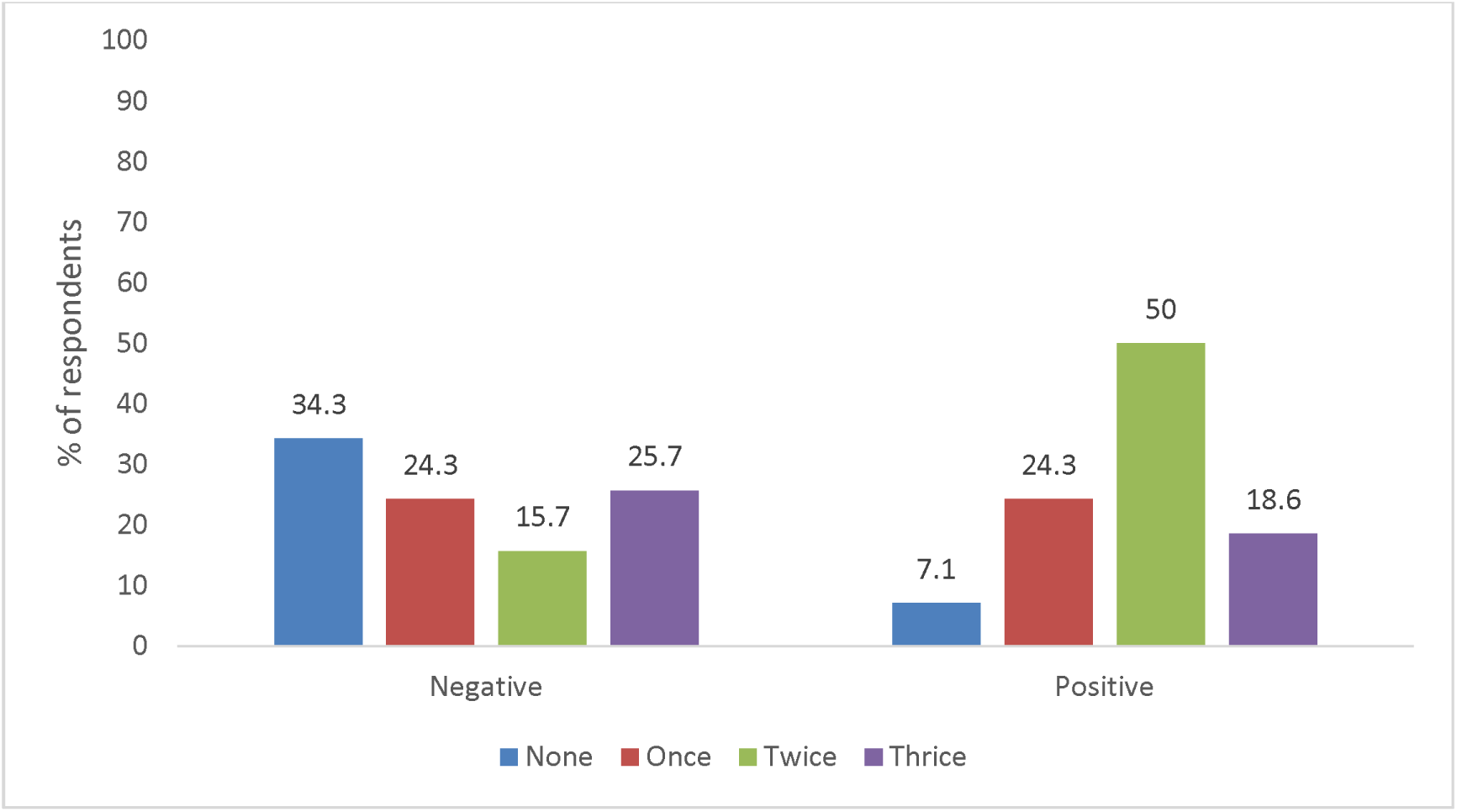
IPT-SP doses received. Percentage illustration of each IPT-SP doses received in both cohorts.

**Supplementary figure 5:**
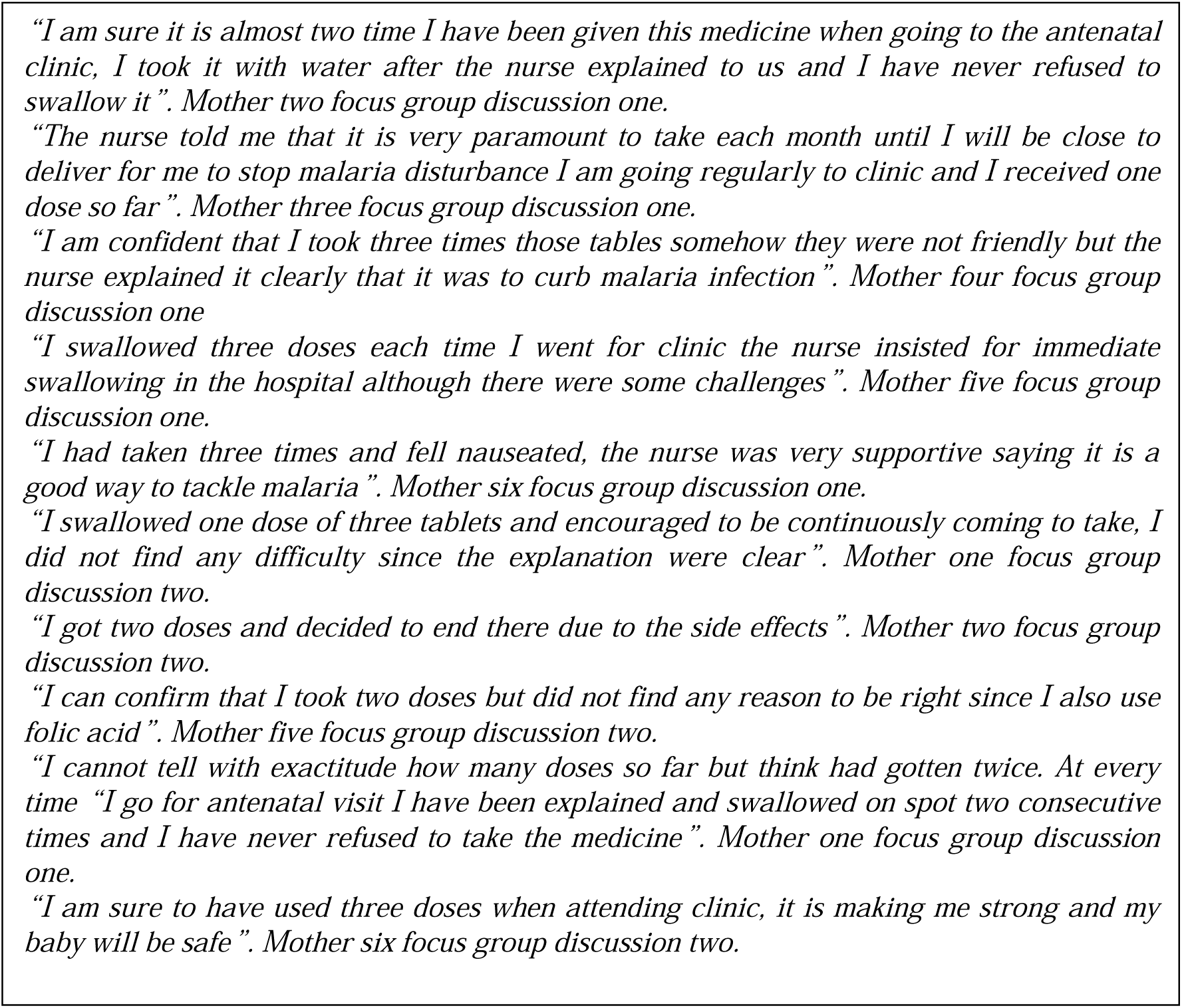
Theme: Practices of intermittent preventive treatment, Sub-theme: Acceptance and completeness of sulfadoxine-pyrimethamine. Focus group discussions illustrating in-depth views using qualitative insight in both cohorts.

## Strengths of the study

The combination of quantitative and qualitative results sound as an appropriate approach to give more insightful considerations on “knowing and doing” aspects of intermittent preventive treatment use in both cohorts.

## Acknowledgement

We acknowledge the collaboration and efforts of Webuye hospital management team in assisting us during all the process of data collection. Kindly find here the expression of our gratitude.

## Abbreviations

ANC: Antenatal clinic
HIV: Human Immunodeficiency Virus
IPT-SP: Intermittent preventive treatment/Sulfadoxine-pyrimethamine
KMS: Kenya Malaria Strategy
LLIN: Long lasting insecticide treated nets
MAXQDA: Computer-assisted qualitative analysis software.
WHO: World Health Organization

